# Subcortical brain volumes in young infants exposed to antenatal maternal depression: Findings from a South African birth cohort

**DOI:** 10.1101/2022.04.21.22273836

**Authors:** Nynke A. Groenewold, Catherine J. Wedderburn, Jennifer A. Pellowski, Jean-Paul Fouché, Liza Michalak, Annerine Roos, Roger P. Woods, Katherine L. Narr, Heather J. Zar, Kirsten A. Donald, Dan J. Stein

## Abstract

**Background:** Several studies have reported enlarged amygdala and smaller hippocampus volumes in children and adolescents exposed to maternal depression. It is unclear whether similar volumetric differences are detectable in the infants’ first weeks of life, following exposure *in utero*. We investigated subcortical volumes in 2-to-6 week old infants exposed to antenatal maternal depression (AMD) from a South African birth cohort.

**Methods:** AMD was measured with the Beck Depression Inventory 2^nd^ edition (BDI-II) at 28-32 weeks gestation. T2-weighted structural images were acquired during natural sleep on a 3T Siemens Allegra scanner. Subcortical regions were segmented based on the University of North Carolina neonatal brain atlas. Volumetric estimates were compared between AMD-exposed (BDI-II⍰20) and unexposed (BDI-II<14) infants, adjusted for age, sex and total intracranial volume using analysis of covariance.

**Results:** Larger volumes were observed in AMD-exposed (*N*=49) compared to unexposed infants (*N*=75) for the right amygdala (1.98% difference, *p*=0.039) and bilateral caudate nucleus (left: 5.78% difference, p=0.001; right: 6.06% difference, *p*<0.001). A significant AMD-by-sex interaction was found for the hippocampus (left: *F*(1,118)=4.80, *p*=0.030; right: *F*(1,118)=5.16, *p*=0.025), reflecting greater volume in AMD-exposed females (left: 5.09% difference, *p*=0.001, right: 3.53% difference, *p*=0.010), but not males.

**Conclusions:** Volumetric differences in subcortical regions can be detected in AMD-exposed infants soon after birth, suggesting structural changes may occur *in utero*. Female infants might exhibit volumetric changes that are not observed in male infants. The potential mechanisms underlying these early volumetric differences, and their significance for long-term child mental health, require further investigation.

## 1. INTRODUCTION

Antenatal maternal depression (AMD) is experienced by approximately 10-15% of pregnant women (Gavin et al., 2005; Woody et al., 2017), and this might be even 2 times higher in low-income settings (Gelaye et al., 2016). Prospective longitudinal studies provide strong evidence that infants who are exposed to maternal depression *in utero* are at increased risk for developing socio-emotional and behavioural problems later in life (O’Donnell et al., 2014; Stein et al., 2014), although evidence from low- and middle-income countries (LMICs) remains scarce (Burger et al., 2020). It has long been hypothesized that physiological processes activated in AMD may impact early neurodevelopment (Field, 1992; Goodman & Gotlib, 1999). Multiple plausible intra-uterine pathways have since been identified, including highly complex and dynamic systems such as those involving glucocorticoid regulation, immunological function, and epigenetic programming (O’Donnell & Meany, 2017). Rapid neural growth and differentiation in the brain of the unborn child may signal a period of increased sensitivity to AMD and other forms of maternal stress (Andersen, 2003), and the hippocampus and amygdala may be particularly susceptible (Adamson et al., 2018). Although it is important for maternal health intervention strategies to understand how early brain development is impacted by AMD, it is unclear at which stage of development structural alterations first emerge in the brain of an exposed child.

Multiple structural imaging studies found evidence for larger amygdala volumes and smaller hippocampal volumes in adolescents exposed to maternal depression earlier in their childhood (Chen et al., 2010; Lupien et al., 2011; Gilliam et al., 2015; also see Pagliaccio et al., 2020 for a null finding in children). Moreover, three birth cohorts have investigated brain structure following AMD exposure. The largest cohort did not detect an association with hippocampus or amygdala volumes in >600 children at 6-10 years of age (El Marroun et al., 2016). For the second cohort, enlarged amygdala volumes were found in 4-year old girls but not boys (total N=203; Wen et al., 2017). In a smaller subsample of 157 neonates, atypical amygdala microstructure was detected in the absence of volumetric differences in this region (Rifkin-Graboi et al., 2013). A small sub-study in the third birth cohort revealed an association between higher AMD symptoms and smaller right amygdala volumes in 4-year old boys but not girls (N=14 in each group; Acosta et al., 2020a). This association was not present in the first two months of life (N=105; Acosta et al., 2020b). However, AMD severity scores indicated mild symptoms in this last cohort. Antenatal exposure to clinically confirmed major depression has been associated with enlarged subcortical volumes in 3-6 months old infants (N=64), but here individual regions were not examined (Sethna et al., 2021). Given the inconsistent findings, much remains unknown regarding infant brain development following AMD exposure.

Depression in pregnant women frequently persists after childbirth (Underwood et al., 2016). The differences in brain morphometry observed in older children exposed to AMD may therefore be confounded by postnatal depression. Moreover, the pattern of these differences may vary as a function of time since exposure and child development (O’Donnell & Meany, 2017). It is therefore critical to examine regional brain volumes in AMD-exposed infants soon after birth, minimizing the effects of postnatal influences. Early neurodevelopmental trajectories differ between the sexes (Buss et al., 2009) and preclinical studies have identified higher glucocorticoid sensitivity and stronger adaptive placental reactions to prenatal stress in female compared to male offspring (Meakin et al., 2021). There is some evidence that regional brain volumes associated with AMD exposure differ as a function of child sex (e.g. Wen et al., 2017; Acosta et al., 2020a), however further investigation is necessary in young infants. Whereas the amygdala and hippocampus have been repeatedly studied in connection with AMD exposure, other subcortical regions linked to depression, including the basal ganglia and thalamus (Bora et al., 2012; Pagliaccio et al., 2020), remain understudied. Finally, the previous studies on regional brain volumes in relation to AMD exposure were conducted in high-income countries. Even though LMICs carry the largest burden of disease from AMD, it is unknown whether AMD exposure impacts early brain development in these settings.

This study aimed to advance understanding of the link between AMD exposure and early brain development using data from a South African longitudinal birth cohort. The aims were: 1) to assess whether enlarged amygdala and smaller hippocampal volumes, as previously observed in children and adolescents after maternal depression exposure, can already be detected in 2-6 week old infants that were exposed to AMD in a LMIC setting; 2) to explore whether additional volumetric differences are present in other subcortical brain regions, in particular in the basal ganglia and thalamus; 3) to examine the hypothesis that regional brain volumes associated with AMD exposure vary as a function of infant sex. Finally, possible associations between the severity of AMD exposure and subcortical volumes were investigated.

## 2. MATERIALS AND METHODS

### 2.1 Study design

The Drakenstein Child Health Study (DCHS) is a longitudinal birth cohort investigating the early life determinants of child health in two low-resource communities located in the Western Cape, South Africa (Zar et al., 2015). Pregnant women were recruited from two public sector clinics for primary health care that serve different populations: Mbekweni (black African community) and TC Newman (mixed ancestry community). The DCHS included women of 18 years and older who attended the recruitment clinics with pregnancy at 20-28 weeks’ gestation and intended to remain in the area. An extensive psychosocial characterization demonstrated a high burden of poverty-related stressors, including low education, substance use and depressive symptoms during pregnancy in this cohort (Stein et al., 2015).

The full DHCS cohort consisted of 1137 expectant mothers who gave birth to 1143 infants (Donald et al., 2018; Zar et al., 2019). Here, data were derived from a nested sub-study of 236 infants (20.6% of full cohort) that underwent brain magnetic resonance imaging (MRI) at 2-6 weeks of age. These infants were selected in a convenience sample that fulfilled age requirements and was enriched for maternal depression exposure. The major exclusion criteria were: i) infant congenital abnormality, genetic syndrome, neurological disorder or HIV infection; ii) neonatal intensive care unit admission; iii) low Apgar score (<7 at 5 minutes); iv) premature birth (<36 weeks gestation); and v) MRI contra-indications, such as ferromagnetic implants.

The DCHS was approved by the University of Cape Town, Faculty of Health Sciences, Human Research Ethics Committee (full cohort: 401/2009; MRI sub-study: 525/2012). Written informed consent was obtained from the mothers on behalf of herself and her infant at enrolment, and again at the start of the neuroimaging session. All study procedures were carried out in accordance with the Declaration of Helsinki (World Medical Association, 2013).

### 2.2 Antenatal maternal depression

Maternal depression was measured with the Beck Depression Inventory II (BDI-II; Beck et al., 1996) at an antenatal study visit between 28 and 32 weeks of gestation. The BDI-II is a well-validated and widely used self-report measure of depressive symptoms (Wang & Gorenstein, 2013; Makhubela & Mashegoane, 2016) that is suitable for perinatal assessments (Bos et al., 2009; Tandon et al., 2012) as conducted in a birth cohort study. The measure consists of 21 items scored from 0 to 3 with increasing severity. A total BDI-II score was obtained through summing all items. The larger DCHS cohort showed a Cronbach’s alpha of 0.90 for the BDI-II and 21.5% prevalence of antenatal maternal depression (AMD: BDI-II ≥20 indicates moderate-to-severe depression according to manual; also see Brittain et al., 2015). In the present investigation, exposed infants (BDI-II ≥20) were compared against infants with minimal exposure to AMD (BDI-II <14 based on manual). Additional antenatal characterization was available from the 10-item Edinburgh Postnatal Depression Scale (EPDS; Cox et al., 1987; Murray & Cox, 1990) at the antenatal study visit and from self-reported medication use at enrolment.

### 2.3 Antenatal maternal substance use

Antenatal exposure to harmful substances may confound the association between infant subcortical brain volumes and antenatal maternal depression (Huizink & De Rooij, 2018). Alcohol use during pregnancy was assessed using a composite measure to counteract potential underreporting of alcohol use. This measure combined the Alcohol, Smoking and Substance Involvement Screening Test (ASSIST; Humeniuk et al., 2008) at 28-32 weeks gestation and two retrospective self-report questionnaires recording hazardous alcohol use in pregnancy (more information in Donald et al., 2019). Alcohol exposure was defined as ASSIST total alcohol score >10 (at least weekly alcohol use with negative consequences) or retrospective self-report of 2 or more alcoholic consumptions per week. Tobacco use was assessed with cotinine measurements in maternal urine using the IMMULITE 1000 Nicotine Metabolite Kit (Siemens Medical Solutions Diagnostics, Glyn Rhonwy, Llanberis, UK). Active smoking was defined by cotinine ≥500 ng/ml (Vanker et al., 2016) in maternal urine collected antenatally or at birth.

### 2.4 Birth characteristics

Gestational age at birth (in weeks) was recorded through ultrasound measurements when available and otherwise was based on measurements of fundal height or self-reported last menstrual period. Birth weight (in kilograms) was obtained at birth at the central hospital where deliveries took place. Maternal age at birth was calculated from maternal and infant dates of birth. All expectant mothers received HIV testing as per national guidelines and infected women were started on antiretroviral therapy if not already on treatment. HIV-exposed infants were tested for HIV using polymerase chain reaction tests at 6 weeks of age (Pellowski et al., 2019). Maternal HIV infection was more frequently recorded in Mbekweni compared to TC Newman (Wedderburn et al., 2022).

### 2.5 Infant MRI scan acquisition and processing

MR images were acquired on a 3T Siemens Magnetom Allegra MRI scanner (Erlangen, Germany) at the Cape Universities Brain Imaging Centre (CUBIC), Tygerberg, Cape Town. Infants were swaddled, fed, and thereafter imaged during natural sleep (without sedation). Earplugs and mini-muffs were used for double ear protection, and the head coil was loaded with a wet clay inlay. Sagittal 3D T2-weighted images were acquired with scan parameters: TR = 3500ms; TE = 354 ms; FOV = 160 × 160 mm; 128 slices; voxel size = 1.3 × 1.3 × 1.0 mm. The sequence took 5 min. 41 s. to acquire. Images were successfully obtained in 183 infants (scan success rate: 77.5%). More information is provided in earlier publications (Donald et al., 2016; Wedderburn et al., 2022).

T2-weighted images were brain-extracted using FSL v5.0. The procedure was repeated to ensure non-brain tissue was adequately removed. Brain images were pre-processed further using Statistical Parametric Mapping software (SPM8) run in Matlab R2017B. Images were first registered and then normalised with modulation to the University of North Carolina neonate T2 template (Shi et al., 2011). Normalised images were segmented into grey matter, white matter and cerebrospinal fluid in accordance with the corresponding neonate probabilistic maps. Alignment to the template and segmentation accuracy was confirmed through visual inspection. GM segmentations from 146 infants passed quality control (flowchart provided in Supplemental Figure A).

The automated anatomical labelling atlas (AAL; Tzourio-Mazoyer et al., 2002) was previously adapted for use in conjunction with the neonate T2 template (Shi et al., 2011). Grey matter volumes were extracted for subcortical regions as defined by this atlas: left and right amygdala, hippocampus, thalamus, caudate, putamen and pallidum. Total grey matter, white matter and cerebrospinal fluid estimates were also extracted and summed to obtain total intracranial volume.

### 2.6 Statistical analyses

Maternal and infant characteristics were compared between AMD-exposed (n=49) and unexposed (n=75) groups with two-sample t-tests or chi-quare tests where appropriate. In the main analysis, volumes of the twelve subcortical regions were compared between AMD-exposed and unexposed infants using one-way analysis of variance including infant sex, age at scan and intracranial volume (ICV) as covariates, taking possible group differences in global brain size into account. Considering the different socio-environmental characteristics associated with clinic site, the main analysis was repeated including clinic site as additional covariate. Furthermore, the analysis was repeated replacing clinic site with maternal age, education level (low: only primary versus high: any secondary education), alcohol exposure and smoking exposure covariates. This last model was estimated to gain insight into specific variables that may confound the identified group differences. Finally, any descriptive variables that demonstrated significant group differences were additionally adjusted for in supplemental analyses. Since depression-related volumetric alterations may differ as a function of child sex, AMD-by-sex interactions were tested for the pre-defined hippocampus and amygdala regions of interest (ROIs), and for other regions that reached significance in the main analysis. For illustrative purposes, analyses of AMD exposure stratified by infant sex were conducted for *a priori* ROIs. Finally, associations between subcortical volumes and depression severity in AMD-exposed infants were examined in a post-hoc analysis.

*Given the a priori* hypotheses for the hippocampus and amygdala, group differences in these ROIs were considered statistically significant at uncorrected p<0.05. For the other eight subcortical regions that were explored, results were evaluated against q<0.05 adjusted for multiple comparisons according to the Benjamini-Hochberg false-discovery rate (FDR) correction (Benjamini & Hochberg, 1995). AMD-by-sex interactions and severity associations were considered significant at uncorrected p<0.05. All statistical analyses were carried out in IBM SPSS v27.

## 3 RESULTS

### 3.1 Sample characteristics for AMD-exposed and unexposed infants

Expectant mothers with AMD experienced moderate-to-severe depressive symptoms (BDI-II M=29.0, SD=7.7). However, only one mother with depressive symptoms reported antidepressant medication use (citalopram) at enrolment. Antenatal tobacco use was more prevalent in the AMD exposed compared to unexposed group (43.8% versus 20.0%, X^2^=7.98, p=0.005), whereas alcohol use was similar across the groups (18.4% and 16.0%, respectively). In this DCHS imaging subsample, AMD exposure differed per clinic site. The majority of expectant mothers with AMD attended TC Newman (67.3%), and the majority of mothers without AMD attended Mbekweni (60.6%; X^2^=8.87, p=0.003). Maternal HIV infection was less prevalent in the AMD-exposed group (12.2% versus 34.7%, X^2^=7.78, p=0.005). Despite the selective inclusion of healthy infants, birth weight was considerably lower in AMD-exposed infants (3.00 vs. 3.29kg, t=3.57, p=0.001). More sample details are provided in Table 1.

**Table 1:** Descriptive characteristics of AMD-exposed and unexposed groups.

| Descriptive variable of interest | AMD-exposed<br>(n=49) | AMD-unexposed<br>(n=75) | Comparison |  |
| --- | --- | --- | --- | --- |
| | M (SD) or n (%) | M (SD) or n (%) | $\chi^2$ or t | p-val. |
| **Infant characteristics** |  |  |  |  |
| Infant sex (% female) | 28 (57.1%) | 35 (46.7%) | 1.30 | .254 |
| Infant age at scan (wks) | 3.03 (0.82) | 3.25 (0.94) | 1.29 | .199 |
| Infant birth weight (kg) | 3.00 (0.41) | 3.29 (0.48) | 3.57 | **.001** |
| Infant birth gestational age (wks) | 38.73 (1.92) | 39.05 (1.87) | 0.92 | .361 |
| Infant intracranial volume (cm<sup>3</sup>) | 426.00 (4.14) | 425.84 (4.89) | -0.20 | .844 |
| **Clinic characteristics** |  |  |  |  |
| Clinic site (% TC Newman) | n=33 (67.3%) | n=30 (40.0%) | 8.87 | **.003** |
| **Maternal characteristics** |  |  |  |  |
| Maternal age at birth (yrs) | 25.94 (5.56) | 27.99 (5.79) | 1.96 | .052 |
| Maternal education level (% low) | n=30 (61.2%) | n=36 (48.0%) | 2.08 | .149 |
| Antenatal alcohol use (% use) | n=9 (18.4%) | n=12 (16.0%) | 0.12 | .731 |
| Antenatal tobacco use (% use)* | n=21 (43.8%) | n=15 (20.0%) | 7.98 | **.005** |
| Antenatal maternal HIV infection | n=6 (12.2%) | n=26 (34.7%) | 7.78 | **.005** |
| Antenatal maternal BDI-II score | 29.02 (7.73) | 4.59 (4.64) | -21.99 | **<.001** |
| Antenatal maternal EPDS score | 14.27 (4.99) | 7.53 (3.63) | -8.69 | **<.001** |

Abbreviations: AMD = antenatal maternal depression; HIV = human immunodeficiency virus; BDI-II = Beck Depression Inventory 2<sup>nd</sup> edition; EPDS = Edinburgh Postnatal Depression Scale. \*Maternal cotinine data unavailable for 1 infant in AMD-exposed group.

### 3.2 Differences in subcortical volumes between AMD-exposed and unexposed infants

The minimally adjusted model, which included infant age at scan, sex and ICV as covariates, revealed volumetric differences in *a priori* ROIs. AMD-exposed infants showed larger bilateral hippocampus volumes (left: mean difference = +5.09%, F(1,119) = 10.58, p = 0.001; right: mean difference = +3.54%, F(1,119) = 6.94, p = 0.010) and right amygdala volumes (mean difference = +1.93%, F(1,119) = 4.36, p = 0.039) compared to unexposed infants. There was no significant group difference for the left amygdala (mean difference = +0.91%, F(1,119) = 1.76, p = 0.187). Adjusted regional volumes are presented in Table 2.

**Table 2:** Differences in subcortical volumes between AMD-exposed and unexposed infants at varying levels of adjustment for potentially confounding factors.

| Subcortical regions | AMD-exposed<br>(n=49) | AMD-unexposed<br>(n=75) | Minimal<br>adjustm. | Minimal +<br>clinic site | Minimal +<br>maternal |
| --- | --- | --- | --- | --- | --- |
| Volumes in mm<sup>3</sup> | M (SE)<sup>1</sup> | M (SE)<sup>1</sup> | p-val.<sup>1</sup> | p-val.<sup>2</sup> | p-val.<sup>3</sup> |
| Regions of Interest |  |  |  |  |  |
| Left Amygdala | 573.34 (3.02) | 568.15 (2.43) | .187 | .059 | .170 |
| Right Amygdala | 515.14 (3.61) | 505.38 (2.91) | .039 | .046 | .089 |
| Left Hippocampus | 1589.07 (18.28) | 1512.10 (14.72) | .001 | .005 | .002 |
| Right Hippocampus | 1673.47 (16.79) | 1616.23 (13.52) | .010 | .024 | .004 |
| Explorative Regions |  |  |  |  |  |
| Left Caudate<sup>5</sup> | 1903.12 (22.79) | 1798.88 (18.35) | .001 | .005 | .001 |
| Right Caudate<sup>6</sup> | 1942.07 (23.41) | 1830.54 (18.86) | <.001 | .005 | <.001 |
| Left Putamen | 2801.59 (12.02) | 2795.53 (9.68) | .697 | .341 | .362 |
| Right Putamen | 2893.27 (14.28) | 2870.24 (11.50) | .215 | .274 | .304 |
| Left Pallidum | 712.88 (4.57) | 711.84 (3.68) | .860 | .489 | .362 |
| Right Pallidum | 712.16 (4.37) | 711.36 (3.52) | .888 | .816 | .340 |
| Left Thalamus | 3194.56 (15.65) | 3161.35 (12.60) | .104 | .118 | .210 |
| Right Thalamus | 3267.48 (14.49) | 3244.54 (11.67) | .224 | .268 | .285 |

<sup>1</sup>Statistics derived from model: AMD (BDI-II $\geq 20$ vs BDI-II $< 14$ ) with infant age at scan, sex, ICV (minimal adjustment)
<sup>2</sup>Statistics derived from model: AMD with infant age at scan, sex, ICV, and clinic site
<sup>3</sup>Statistics derived from model: AMD with infant age at scan, sex, ICV, and maternal age, education, tobacco, alcohol use
<sup>5</sup>The False Discovery Rate corrected threshold for significance in the left caudate nucleus is $p < 0.01250$
<sup>6</sup>The False Discovery Rate corrected threshold for significance in the right caudate nucleus is $p < 0.00625$

After additional adjustment for clinic site, similar findings were recorded (left hippocampus: mean difference = +4.66%, p = 0.005; right hippocampus: mean difference = +3.18%, p = 0.024; right amygdala: mean difference = +1.94%, p = 0.046). After full adjustment for maternal characteristics, the hippocampal differences slightly gained in magnitude (left hippocampus: mean difference = +5.31%, p = 0.002; right hippocampus: mean difference = +4.20%, p = 0.004), whereas significance was lost for the right amygdala (mean difference = +1.69%, p = 0.089). Of note, none of the added maternal characteristics were significantly associated with right amygdala volume (all *p* > 0.31). Two sensitivity analyses separately excluding infants exposed to maternal HIV infection and infants born late preterm (<37 weeks gestation) and a supplemental analysis including birth weight as additional covariate demonstrated robustness of effects (Supplemental Table A1/2), although significance was lost for volume of the right amygdala in the preterm birth and birth weight subanalyses.

The exploratory analyses revealed enlarged bilateral caudate volumes in AMD-exposed compared to unexposed infants. These group differences were significant after multiple comparison correction in the minimally adjusted model (left caudate: mean difference = +5.79%, F(1,119) = 12.49, p = 0.001; right caudate: mean difference = +6.09%, F(1,119) = 13.55, p < 0.001). Moreover, the volumetric differences remained significant after adjustment for clinic site (left caudate: mean difference = +4.80%, p = 0.005; right caudate: mean difference = +4.83%, p = 0.005) and maternal characteristics (left caudate: mean difference = +5.90%, p = 0.001; right caudate: mean difference = +6.35%, p < 0.001). The thalamus, pallidum and putamen did not show significant group differences in volume at any level of adjustment (all *p* > 0.10; Table 2). Subcortical regions with significant group differences in volume are depicted in Figure 1.

**Figure 1:**
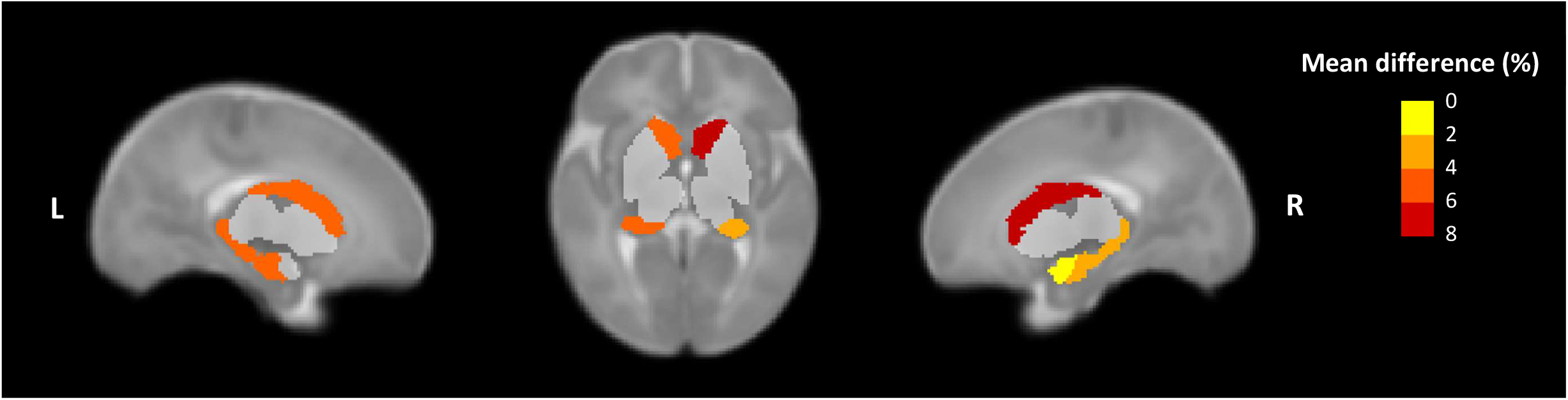
Percentage mean difference in grey matter volume for the subcortical regions that showed significant differences between AMD-exposed and AMD-unexposed infants* after adjusting for infant age, sex and intracranial volume, visualized in a neonate template brain. * The subcortical regions that did not show a significant group difference in volume are included for anatomical reference, in pale grey.

### 3.3 Volumetric differences associated with AMD exposure in female and male infants

A significant AMD-by-sex interaction was observed bilaterally for the hippocampus (left: F(1,118) = 4.80, p=0.030; right: F(1,118) = 5.16, p = 0.025). Hereafter, analyses stratified by sex demonstrated a significant enlargement in AMD-exposed compared to unexposed female infants (left hippocampus: mean difference = +8.76%, F(1,59) = 15.22, p = 0.001; right hippocampus: mean difference = +6.47%, F(1,59) = 12.73, p = 0.010). No significant differences in hippocampal volume were found in male infants (left: p = 0.576 and right: p = 0.882). Moreover, no significant AMD-by-sex interactions were detected for the amygdala or caudate (all *p* > 0.28; see Supplemental Table B). Figure 2 presents the volumes of *a priori* ROIs according to AMD exposure groups, separately for female and male infants.

**Figure 2:**
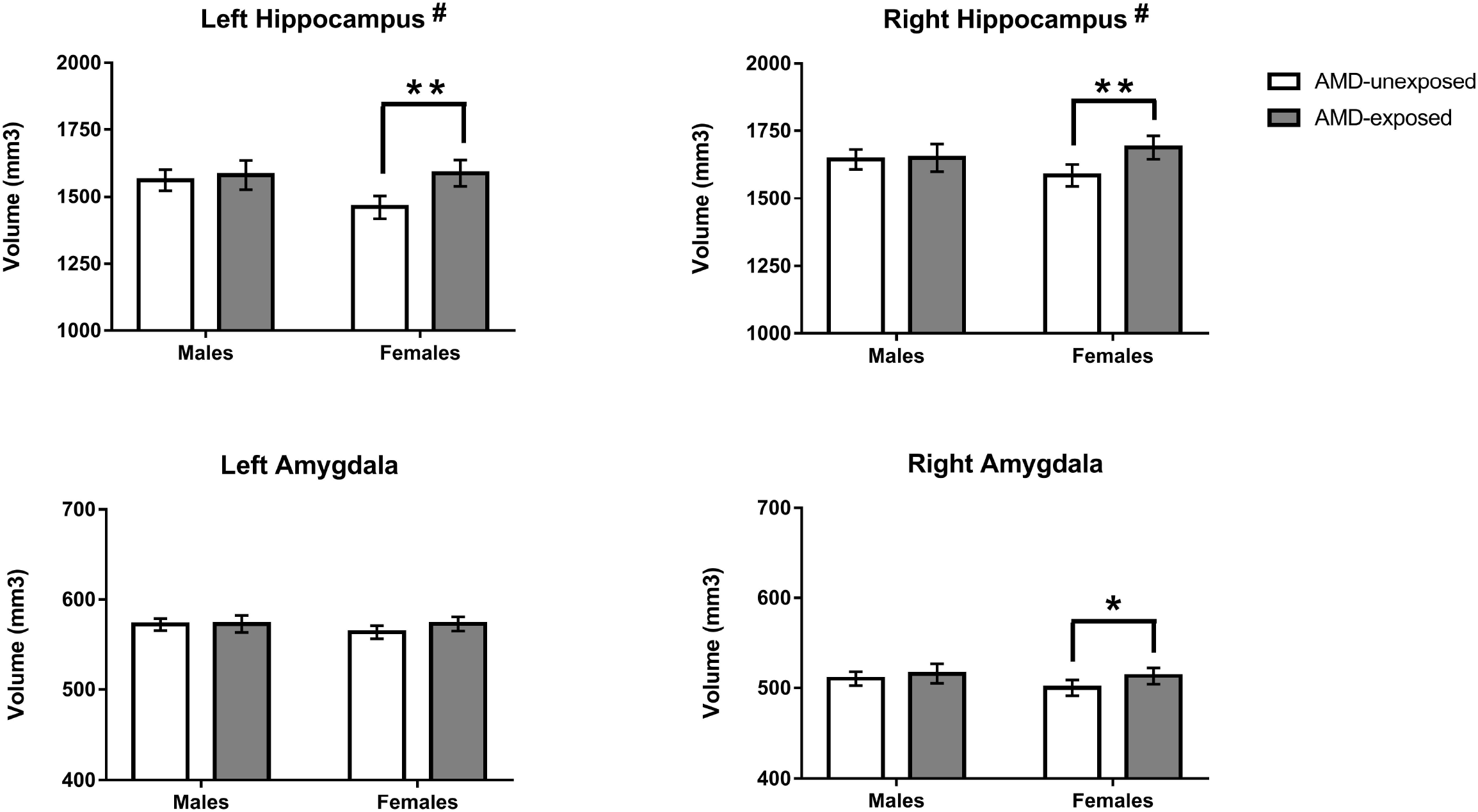
Differences in hippocampus and amygdala volume related to AMO exposure presented separately for female and male infants. # Subcortical region with a significant AMO-by-sex interaction (p<0.05) * Significant difference between AMO-exposed and AMO-unexposed female infants (p<0.05) ** Significant difference between AMO-exposed and AMO-unexposed female infants (p<0.01)

### 3.4 The association between AMD severity and subcortical volumes in AMD-exposed infants

No significant associations between AMD severity and subcortical volumes were detected in AMD-exposed infants for the BDI-II (all p ≥ 0.093) in regression models adjusted for sex, age and ICV. EPDS scores showed a trend-level association with right amygdala volume (β = 0.284, p = 0.054).

## 4. DISCUSSION

This study in South African infants aged 2-6 weeks detected enlarged right amygdala and bilateral hippocampus volumes in AMD-exposed compared to unexposed infants. In an exploratory analysis of other subcortical regions, robust evidence was found for an enlargement of the bilateral caudate nucleus in AMD-exposed infants. The direction of effect for the hippocampus was unexpected, in light of the smaller volumes previously observed after maternal depression exposure in childhood. As hypothesized, the regional brain volumes associated with AMD exposure varied as a function of infant sex: enlarged hippocampal volumes were apparent in females but not males exposed to AMD. However, no significant interactions with infant sex were observed for the amygdala and caudate nucleus. Furthermore, no significant associations between severity of AMD exposure and subcortical volumes were found. Taken together, the findings confirm that volumetric differences in subcortical regions can be detected in AMD-exposed infants soon after birth in a LMIC setting.

The findings of enlarged total grey matter volumes of *a priori* amygdala and hippocampus ROIs build on findings from two previous birth cohort studies. The first study reported an atypical amygdala microstructure that was most pronounced in the right amygdala for AMD-exposed compared to unexposed neonates (Rifkin-Graboi et al., 2013). Consistent with the present study, no volumetric differences were identified after full adjustment for exposure and birth variables. The second study also did not find a main effect of AMD exposure, but did report a gene-by-environment interaction for right amygdala volume, with a weak positive association apparent in infants at low genetic risk of developing depression (Acosta et al., 2020b; partial replication of Qiu et al., 2017). Of note, the larger volume of the right amygdala identified in our study did not retain significance in the fully adjusted model, which was mostly attributable to adjustment for birth weight. Low birth weight or impaired intrauterine growth is more likely to be the consequence rather than the cause of AMD, and adjustment for birth weight might mask true group differences. The loss of significance might be indicative of a mediating physiological mechanism, such as excessive intra-uterine glucocorticoid exposure, that impacts both foetal growth and neurodevelopment (O’Donnell & Meany, 2017). Even though confounding by genetic factors cannot be ruled out, the results from the three birth cohorts support the hypothesis that intra-uterine pathways are likely to play a significant role in alterations in brain structure following AMD exposure, especially for structural alterations in the right amygdala.

When comparing the present findings to volumetric differences observed in older children exposed to maternal depression, a mostly consistent direction of effect is observed for the amygdala but not the hippocampus (Lupien et al., 2011; Wen et al., 2017; Chen et al., 2019). It is possible that the time of exposure impacts volumetric differences in such a way that larger hippocampal volumes follow from antenatal exposure, and smaller hippocampal volumes follow from postnatal exposure. However, it is more plausible that volumetric differences are not static but change as a function of brain maturation and time since exposure (in line with the slowed hippocampal growth previously reported for infants exposed to antenatal maternal anxiety; Qiu et al., 2013). In our cohort, greater volume of the hippocampus was evident in female but not male AMD-exposed infants. This is consistent with preclinical observations of alterations in hippocampal structure in female but not male offspring after prenatal stress (e.g. Zhu et al., 2004; Behan et al., 2011; Bock et al., 2011). These studies reported neuronal loss, glial deficits and loss of dendritic complexity in pre-pubertal female rodents. However, neuronal loss is a gradual process (Zhu et al., 2004), and it remains unclear to what extent such structural deficits in the hippocampus occur in the newborn period (Weinstock, 2011). More basic and clinical research is needed to gain insight into the developmental trajectories of amygdala and hippocampus volumes in male and female children exposed to AMD, and their association with the subsequent onset of child psychopathology.

In addition to the *a priori* ROIs, the caudate nucleus was newly implicated in the context of AMD exposure. Whereas the caudate is not the most consistently implicated subcortical brain region in the neurobiology of depression, meta-analyses of adult depressed patients (Arnone et al., 2012; Bora et al., 2012) and two studies in depressed adolescents (Matsuo et al., 2008; Shad et al., 2012) have identified smaller caudate volumes. The caudate nucleus is an important region in the brain reward network and is thought to play a role in anhedonic depressive symptoms (Enneking et al., 2019). A recent neuroimaging investigation identified a positive association between polygenic risk for depression and caudate volumes in male neonates, as well as a negative association between polygenic risk for depression in female neonates (Acosta et al., 2020c). Here, we report enlarged caudate volumes in male and female infants exposed to AMD. In light of the findings from Acosta and colleagues (2020c), there may be a genetic component to this association. Indeed, bilateral enlargement in caudate volume after AMD exposure was robust against adjustment for birth weight, maternal HIV and substance use exposure. However, given that our study is the first to link AMD exposure to caudate volume, there is a need for replication.

Three subcortical brain regions were found to have a larger volume in AMD-exposed infants 2-6 weeks after birth, when taking total intracranial volume into account. The consistent direction of effect at this developmental stage, as well as the slight attenuation of effect size when adjusting for antenatal and birth variables, suggests that specifically antenatal exposure to AMD influences early brain development. Genetic risk for depression would be less likely to explain the complete set of findings, given the inconsistent direction of effects compared to volumetric differences in depressed patients and individuals with a family history of depression (including postnatal maternal depression; see Jacobs et al., 2015) and also considering the unique epidemiology of antenatal depression. AMD is highly prevalent compared to depression in other life stages, especially in LMIC settings (Gelaye et al., 2016). There are multiple plausible neurobiological pathways for enlarged infant subcortical volumes after AMD related to epigenetic programming, immunological function, and glucocorticoid regulation. Of special interest are the effects that AMD can have on lowering expression of placental 11β-HSD2, which consequently increases exposure of the foetus to maternal cortisol (O’Donnell & Meany, 2017). Given the stage of neurodevelopment (Andersen, 2003), several processes could be disrupted; prolonged proliferation and differentiation and delayed or reduced apoptosis may occur in the affected subcortical regions. As such, the present study adds to the growing recognition that antenatal maternal depression can impact neurodevelopment during pregnancy and underscores the potential for intervention in pregnancy to benefit both mother and child.

The present study was characterized by several notable strengths. The DCHS is a longitudinal birth cohort with extensive psychosocial characterization and this allowed us to rigorously adjust our group comparisons for possible confounding variables, including maternal substance use. MRI scans were obtained in young infants at 2-6 weeks of age, limiting the confounding effects of unmeasured postnatal exposures. Finally, we present the first investigation of early brain development after AMD exposure in a LMIC population. The relatively high prevalence of clinically relevant AMD symptoms and also of poverty-related stressors (also see Herba et al., 2016) in the communities from which pregnant women were recruited may have enhanced our sensitivity to detect volumetric differences in subcortical brain regions.

However, our findings need to be interpreted with recognition of the study limitations. AMD was measured at a single timepoint in late pregnancy and therefore findings may not generalize to AMD exposure earlier in pregnancy. Mild overestimation of BDI-II severity scores may occur due to somatic symptoms associated with pregnancy, however inclusion of somatic symptoms is critically important for construct validity (Manian et al., 2014). Medication use was recorded in the DCHS cohort through self-report at enrolment and no information was available about medication use later in pregnancy. However, only one pregnant woman reported antidepressant use (citalopram) at enrolment, in line with the very limited access to psychiatric services in underserved communities in the South African population (Seedat et al., 2008). The present study used a lenient definition of the exclusion criterion preterm birth (1 week below the standard definition of 37 weeks gestation; WHO, 1997) to account for the relatively high frequency of preterm births in South Africa (Chawanpaiboon et al., 2019; Jeena et al., 2020). While early brain development following AMD exposure in children born late preterm is epidemiologically relevant, especially in LMIC settings, sensitivity analyses were conducted to ensure the findings also applied to term infants. Due to limited contrast between grey and white matter in the T2-weighted images related to low myelination, we were unable to perform more fine-grained analyses of subcortical volumetric differences.

In conclusion, greater volumes of subcortical brain regions were detected in South African AMD-exposed compared to unexposed infants soon after birth, suggesting structural changes may occur *in utero*. These volumetric differences were most pronounced in female infants, especially for the hippocampus, and this could be indicative of an increased sensitivity to the effects of stress on the intra-uterine environment in female infants. The functional significance of the structural differences remains to be determined, most importantly their predictive value for subsequent problems in child mental health. More research is needed to delineate the neurodevelopmental trajectories of volumetric differences in subcortical brain regions after AMD exposure, and their underlying mechanisms, in LMIC settings.

## Data Availability

The Drakenstein Child Health Study is committed to the principle of data sharing. Deidentified data will be made available to requesting researchers as appropriate. Requests for collaborations to undertake data analysis are welcome. More information can be found on our website [http://www.paediatrics.uct.ac.za/scah/dclhs].

## ACKNOWLEDGEMENTS

We first and foremost thank the families and children who participated in this study. We recognise the study staff at Mbekweni and T.C. Newman clinics, the clinical and administrative staff at Paarl Hospital, and the radiographers at the Cape Universities Brain Imaging Centre at Tygerberg Hospital for their support of the study.

