## Supplemental Materials for "Subcortical brain volumes in young infants exposed to antenatal maternal depression: Findings from a South African birth cohort"

**Supplemental Table A.1:** Post-hoc sensitivity analysis examining the robustness of volumetric differences associated with antenatal maternal depression (AMD) exposure in the amygdala, hippocampus and caudate nucleus, after excluding infants that were exposed to maternal Human Immunodeficiency Virus (HIV) infection and after excluding infants with late preterm birth (PTB).

| **Subcortical region** | **Reference AMD**  ***F*-val.^1^** | **Reference**  **AMD**  ***p*-val.^1^** | **Excl. HIV**  **AMD**  ***F*-val.^2^** | **Excl. HIV**  **AMD**  ***p*-val.^2^** | **Excl. PTB**  **AMD**  ***F*-val.^2^** | **Excl. PTB**  **AMD**  ***p*-val.^2^** |
| --- | --- | --- | --- | --- | --- | --- |
| Left Amygdala | 1.763 | .187 | 1.550 | .216 | 1.120 | .292 |
| Right Amygdala | 4.360 | **.039** | 4.134 | **.045** | 2.265 | .135 |
| Left Hippocampus | 10.584 | **.001** | 10.804 | **.001** | 7.544 | **.007** |
| Right Hippocampus | 6.941 | **.010** | 7.925 | **.006** | 5.227 | **.024** |
| Left Caudate | 12.494 | **.001** | 15.274 | **<.001** | 12.683 | **.001** |
| Right Caudate | 13.549 | **<.001** | 9.804 | **.002** | 12.082 | **.001** |

^1^Statistics derived from minimally adjusted model: AMD (BDI II ≥20 vs BDI-II <14) with infant age at scan, sex, ICV

^2^Statistics derived from minimally adjusted model, after exclusion of 32 infants exposed to maternal HIV infection

^3^Statistics derived from minimally adjusted model, after exclusion of 14 infants born preterm (<37 weeks gestation)

**Supplemental Table A.2:** Post-hoc analysis examining the robustness of volumetric differences associated with antenatal maternal depression (AMD) exposure in the amygdala, hippocampus and caudate nucleus when including infant birth weight (BW) as additional covariate in the fully adjusted model.

| **Subcortical region** | **Reference AMD**  ***F*-val.^1^** | **Reference**  **AMD**  ***p*-val.^1^** | **Post-hoc**  **AMD**  ***F*-val.^2^** | **Post-hoc**  **AMD**  ***p*-val.^2^** | **Post-hoc**  **BW**  ***F*-val.^2^** | **Post-hoc**  **BW**  ***p*-val.^2^** |
| --- | --- | --- | --- | --- | --- | --- |
| Left Amygdala | 1.910 | .170 | .295 | .588 | 11.091 | **.001** |
| Right Amygdala | 2.944 | .089 | 1.041 | .310 | 7.144 | **.009** |
| Left Hippocampus | 9.943 | **.002** | 6.253 | **.014** | 5.710 | **.019** |
| Right Hippocampus | 8.503 | **.004** | 4.723 | **.032** | 8.267 | **.005** |
| Left Caudate | 11.018 | **.001** | 14.732 | **<.001** | 5.030 | **.027** |
| Right Caudate | 12.870 | **<.001** | 14.976 | **<.001** | 2.242 | .137 |

^1^Statistics derived from model: AMD (BDI II ≥20 vs BDI-II <14) with infant age at scan, sex, ICV, maternal age, education, tobacco, alcohol use

^2^Statistics derived from model: AMD (BDI II ≥20 vs BDI-II <14) with infant age at scan, sex, ICV, maternal age, education, tobacco, alcohol use, and infant birth weight

**Supplemental Table B:** Interactions between antenatal maternal depression (AMD) exposure and biological sex of the infant associated with amygdala, hippocampus and caudate nucleus volumes.

| **Subcortical region** | **AMD-by-sex^1^**  ***p*-val.** | **AMD^2^ mean difference** | **AMD^2^**  ***F*-val.^3^** | **AMD^2^**  ***p*-val.** |
| --- | --- | --- | --- | --- |
| Left amygdala  All infants  Males^4^  Females^5^ | .281 | +0.91%  +0.13%  +1.62% | 1.763  .000  2.806 | .187  .989  .099 |
| Right amygdala  All infants  Males^4^  Females^5^ | .402 | +1.93%  +1.10%  +2.69% | 4.360  .439  5.062 | **.039**  .510  **.028** |
| Left hippocampus  All infants  Males^4^  Females^5^ | **.030** | +5.09%  +1.22%  +8.76% | 10.584  .317  15.224 | **.001**  .576  **<.001** |
| Right hippocampus  All infants  Males^4^  Females^5^ | **.025** | +3.54%  +0.37%  +6.47% | 6.941  .022  12.730 | **.010**  .882  **.001** |
| Left caudate  All infants  Males^4^  Females^5^ | .432 | +5.79%  +4.35%  +7.13% | 12.494  4.144  8.739 | **.001**  **.046**  **.004** |
| Right caudate  All infants  Males^4^  Females^5^ | .416 | +6.09%  +4.61%  +7.46% | 13.549  4.393  10.247 | **<.001**  **.041**  **.002** |

^1^Statistics derived from model: AMD (BDI II ≥20 vs BDI-II <14) with infant age at scan, sex, ICV, and AMD-by-sex

^2^Statistics derived from model: AMD (BDI II ≥20 vs BDI-II <14) with infant age at scan, sex, ICV

^4^Sample composition: N=28 AMD-exposed female infants and N=35 AMD-unexposed female infants

^5^Sample composition: N=21 AMD-exposed male infants and N=40 AMD-unexposed male infants

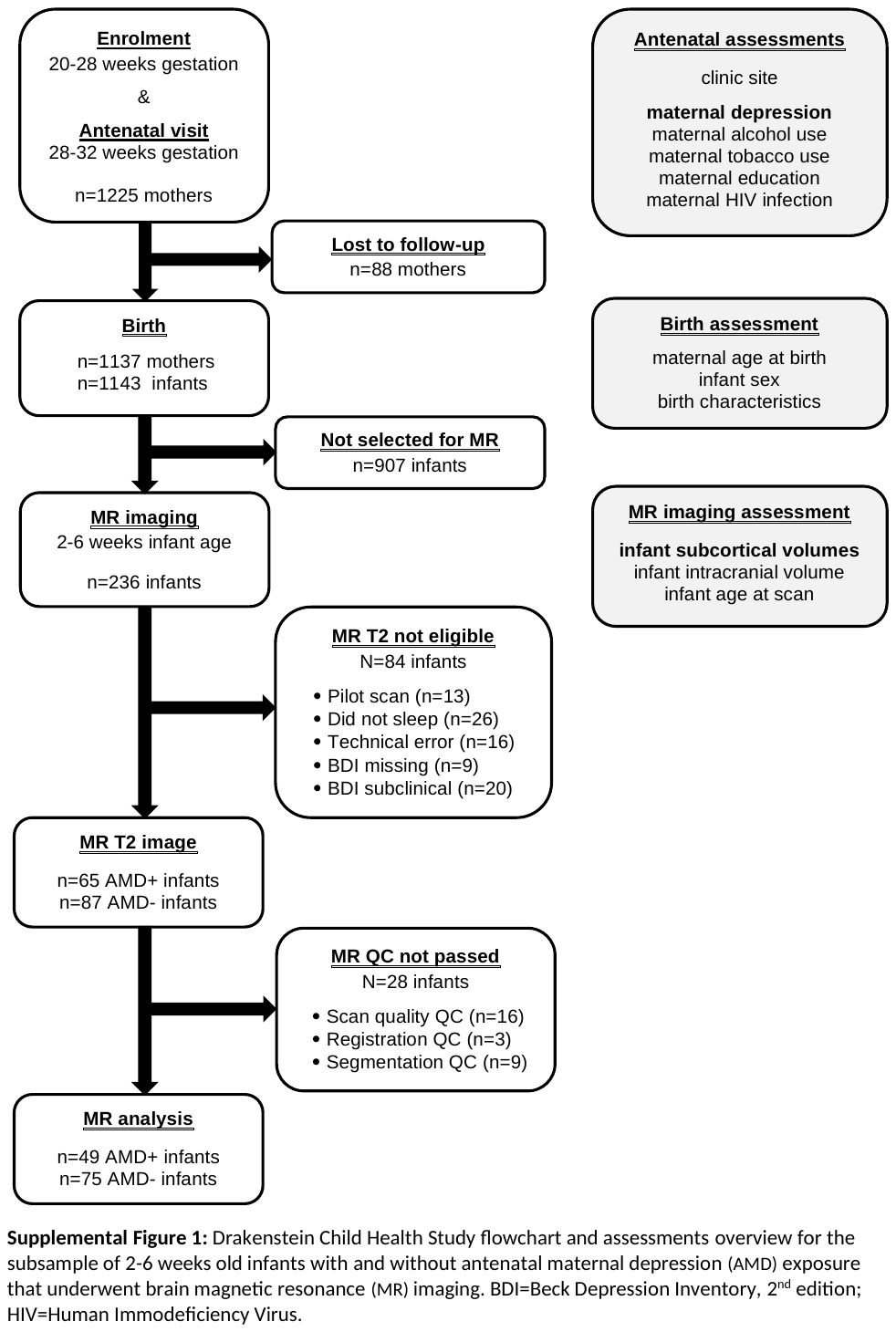
